# Circulating PACAP levels are associated with increased amygdala-default mode network resting-state connectivity in posttraumatic stress disorder

**DOI:** 10.1101/2023.02.28.23286457

**Authors:** KJ Clancy, Q Devignes, P Kumar, V May, SE Hammack, E Akman, EJ Casteen, CD Pernia, SA Jobson, MW Lewis, NP Daskalakis, WA Carlezon, KJ Ressler, SL Rauch, IM Rosso

**Affiliations:** Division of Depression and Anxiety Disorders, McLean Hospital, Belmont, MA; Department of Psychiatry, Harvard Medical School, Boston, MA; Larner College of Medicine, University of Vermont, Burlington, VT; Stanley Center for Psychiatric Research, Broad Institute of MIT and Harvard, Cambridge, MA

**Keywords:** posttraumatic stress disorder, resting-state functional connectivity, default mode network, amygdala, pituitary adenylate cyclase-activating polypeptide

## Abstract

The pituitary adenylate cyclase-activating polypeptide (PACAP) system is implicated in posttraumatic stress disorder (PTSD) and related amygdala-mediated arousal and threat reactivity. PTSD is characterized by increased amygdala reactivity to threat and, more recently, aberrant intrinsic connectivity of the amygdala with large-scale resting state networks, specifically the default mode network (DMN). While the influence of PACAP on amygdala reactivity has been described, its association with intrinsic amygdala connectivity remains unknown. To fill this gap, we examined functional connectivity of resting-state functional magnetic resonance imaging (fMRI) in eighty-nine trauma-exposed adults (69 female) screened for PTSD symptoms to examine the association between blood-borne (circulating) PACAP levels and amygdala-DMN connectivity. Higher circulating PACAP levels were associated with increased amygdala connectivity with posterior DMN regions, including the posterior cingulate cortex/precuneus (PCC/Precun) and left angular gyrus (lANG). Consistent with prior work, this effect was seen in female, but not male, participants and the centromedial, but not basolateral, subregions of the amygdala. Clinical association analyses linked amygdala-PCC/Precun connectivity to anxious arousal symptoms, specifically exaggerated startle response. Taken together, our findings converge with previously demonstrated effects of PACAP on amygdala activity in PTSD-related processes and offer novel evidence for an association between PACAP and intrinsic amygdala connectivity patterns in PTSD. Moreover, these data provide preliminary evidence to motivate future work ascertaining the sex- and subregion-specificity of these effects. Such findings may enable novel mechanistic insights into neural circuit dysfunction in PTSD and how the PACAP system confers risk through a disruption of intrinsic resting-state network dynamics.

## INTRODUCTION

Posttraumatic stress disorder (PTSD) is a severe psychiatric disorder that is characterized, in part, by sustained states of arousal and threat reactivity following a traumatic experience. PTSD is a leading cause of global disease burden and has systemic influences on emotional, neural, and broader physical health through long-lasting alterations of biological stress and arousal systems [1–3]. As such, multilevel systems regulating stress-mediated arousal have been implicated in vulnerability and etiopathology models of PTSD and provide key targets for investigating risk and disease progression factors [4,5].

The pituitary adenylate cyclase-activating polypeptide (PACAP) system regulates stress responsiveness and physiological arousal and has increasingly gained recognition in PTSD. PACAP is a highly-conserved neuropeptide across species that modulates the stress response through its peripheral and central signaling in the hypothalamic-pituitary-adrenal (HPA) axis and autonomic stress pathways [6–8]. Paralleling decades of well-documented dysfunction of the HPA axis in PTSD [9,10], higher circulating blood levels of long-form PACAP (PACAP38) have been found in individuals with (vs. without) PTSD, especially in participants assigned female at birth [11]. Moreover, blood-borne (circulating) PACAP and allelic variations of the predominant PACAP receptor (PAC1R) gene (*ADCYAP1R1*) are associated with PTSD symptom severity, specifically hyperarousal symptoms in female patients [11–14]. As such, PACAP systems may contribute to PTSD through their actions on stress-mediated arousal processes, and this pathway may be sex-dependent.

PACAP and PAC1R are densely expressed within the amygdala complex – the hub of a canonical threat circuit heavily implicated in the neuroanatomy of PTSD and related states of arousal and threat reactivity [15–18]. Prior work has linked PTSD-related polymorphisms of the PAC1R gene to increased amygdala reactivity to threat in humans [19] and to upregulation of PAC1R gene expression in the amygdala following fear conditioning in rodents [11]. Further evidence suggests this PACAP-PAC1R expression and signaling may be preferential to the central (CeA), versus basolateral (BLA), nucleus of the amygdala, and the adjacent, densely-connected bed nucleus of the *stria terminalis* (BNST) which together form the central extended amygdala [7,8,20,21]. Consistent with the association between PACAP and symptoms of (hyper)arousal in PTSD, the CeA is integral to the expression of ‘fear’ and ‘panic’ reflexes in response to threat through axonal projections to associated arousal systems [22]. As such, preclinical studies have demonstrated a direct influence of PACAP on CeA activity and CeA-mediated fear responding [20,23–25]. Taken together, these findings emphasize the potential role of PACAP on threat reactivity in stress/trauma-related disorders through the modulation of amygdala activity, which may be specific to the CeA.

While PACAP effects on amygdala signaling and activity have been well-studied in the context of threat reactivity, little is known about how the peptide influences intrinsic amygdala connectivity, especially in humans. Recently, PTSD has been increasingly characterized by a disrupted equilibrium of intrinsic resting-state neural networks [26–28]. Specific emphasis has been placed on aberrant coupling between salience network (SN) hubs, responsible for bottom-up arousal and detection of threat, and the default mode network (DMN), responsible for homeostatic coordination of large-scale neural activity at rest. Notably, the amygdala is a central hub of the SN whose exaggerated coupling with the DMN at rest has been implicated in both stress-induced hyperarousal and PTSD [27,29–33], and this connectivity pattern may also be sex-dependent [34]. Because these intrinsic brain networks are increasingly viewed as viable mechanistic links between lower-order risk markers (i.e., genetics) and higher-order processes (i.e., task-response, behavior) [35–40], mechanistic accounts for such aberrant amygdala connectivity profiles may yield critical insights into biologically-based vulnerability and etiopathology models of PTSD.

Expanding upon the extant literature demonstrating PACAP modulation of amygdala activity, we sought to characterize the association between circulating PACAP levels and intrinsic amygdala connectivity in PTSD. Here we utilized resting-state functional connectivity of functional magnetic resonance imaging (fMRI) data in trauma-exposed adults screened for PTSD symptoms to test the hypothesis that elevated serum levels of PACAP would be associated with exaggerated amygdala connectivity with the DMN. Moreover, in keeping with the extant literature suggesting a specificity of PACAP effects to participants assigned female (vs. male) at birth and the CeA (vs. BLA), we tested the sensitivity of these associations to sex and amygdala subregions. Finally, clinical associations of identified PACAP-related connectivity profiles with relevant symptoms of hyperarousal and threat reactivity were assessed.

## MATERIALS AND METHODS

### Participants

One-hundred and thirty-six (136) trauma-exposed adults were recruited and enrolled via advertisements in the local community. Study procedures were approved by the Mass General Brigham Human Research Committee and all participants provided written informed consent. Participants were included if they met diagnostic criteria for PTSD based on the Clinician Administered PTSD Scale for DSM-5 (CAPS-5) or if they had subthreshold symptoms, defined as meeting diagnostic criteria for at least two symptom clusters [41]. Given our interest in sex differences motivated by prior work [42], participants were required to be the same sex as assigned at birth, female participants were to be premenopausal, and participants with a history of receiving hormonal replacement therapy or undergoing surgery to change biological sex were excluded. Other exclusion criteria are detailed in the Supplemental Methods.

The current sample consisted of 113 participants with resting-state fMRI data, of which 93 had usable data (excluded: excessive motion = 7, incomplete scanning = 3, inadequate structural-functional alignment = 3, significant artifact based on visual inspection = 7). Of these 93 participants, 89 had detectable serum PACAP levels, resulting in the final analyzed sample cohort of n = 89. Demographic and clinical details are provided in **Table 1**.

**Table 1.**
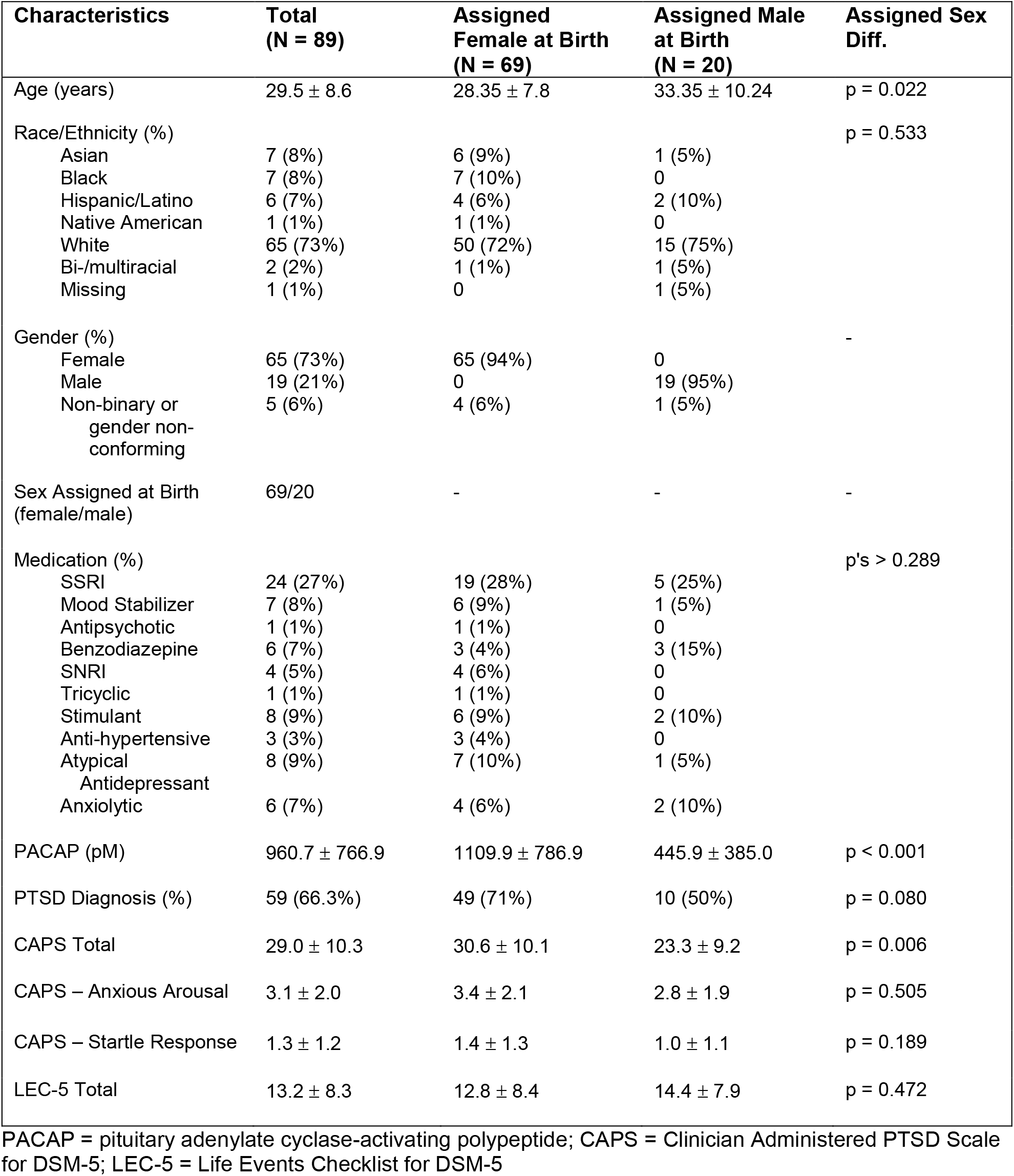
Demographic and clinical characteristics of the sample. Means ± standard deviations or N (%).

| Characteristics | Total<br>(N = 89) | Assigned<br>Female at Birth<br>(N = 69) | Assigned Male<br>at Birth<br>(N = 20) | Assigned Sex<br>Diff. |
| --- | --- | --- | --- | --- |
| Age (years) | 29.5 ± 8.6 | 28.35 ± 7.8 | 33.35 ± 10.24 | p = 0.022 |
| Race/Ethnicity (%) |  |  |  | p = 0.533 |
| Asian | 7 (8%) | 6 (9%) | 1 (5%) |  |
| Black | 7 (8%) | 7 (10%) | 0 |  |
| Hispanic/Latino | 6 (7%) | 4 (6%) | 2 (10%) |  |
| Native American | 1 (1%) | 1 (1%) | 0 |  |
| White | 65 (73%) | 50 (72%) | 15 (75%) |  |
| Bi-/multiracial | 2 (2%) | 1 (1%) | 1 (5%) |  |
| Missing | 1 (1%) | 0 | 1 (5%) |  |
| Gender (%) |  |  |  | - |
| Female | 65 (73%) | 65 (94%) | 0 |  |
| Male | 19 (21%) | 0 | 19 (95%) |  |
| Non-binary or<br>gender non-<br>conforming | 5 (6%) | 4 (6%) | 1 (5%) |  |
| Sex Assigned at Birth<br>(female/male) | 69/20 | - | - | - |
| Medication (%) |  |  |  | p's > 0.289 |
| SSRI | 24 (27%) | 19 (28%) | 5 (25%) |  |
| Mood Stabilizer | 7 (8%) | 6 (9%) | 1 (5%) |  |
| Antipsychotic | 1 (1%) | 1 (1%) | 0 |  |
| Benzodiazepine | 6 (7%) | 3 (4%) | 3 (15%) |  |
| SNRI | 4 (5%) | 4 (6%) | 0 |  |
| Tricyclic | 1 (1%) | 1 (1%) | 0 |  |
| Stimulant | 8 (9%) | 6 (9%) | 2 (10%) |  |
| Anti-hypertensive | 3 (3%) | 3 (4%) | 0 |  |
| Atypical<br>Antidepressant | 8 (9%) | 7 (10%) | 1 (5%) |  |
| Anxiolytic | 6 (7%) | 4 (6%) | 2 (10%) |  |
| PACAP (pM) | 960.7 ± 766.9 | 1109.9 ± 786.9 | 445.9 ± 385.0 | p < 0.001 |
| PTSD Diagnosis (%) | 59 (66.3%) | 49 (71%) | 10 (50%) | p = 0.080 |
| CAPS Total | 29.0 ± 10.3 | 30.6 ± 10.1 | 23.3 ± 9.2 | p = 0.006 |
| CAPS – Anxious Arousal | 3.1 ± 2.0 | 3.4 ± 2.1 | 2.8 ± 1.9 | p = 0.505 |
| CAPS – Startle Response | 1.3 ± 1.2 | 1.4 ± 1.3 | 1.0 ± 1.1 | p = 0.189 |
| LEC-5 Total | 13.2 ± 8.3 | 12.8 ± 8.4 | 14.4 ± 7.9 | p = 0.472 |

PACAP = pituitary adenylate cyclase-activating polypeptide; CAPS = Clinician Administered PTSD Scale for DSM-5; LEC-5 = Life Events Checklist for DSM-5

Participants completed a fasting blood draw, clinical interview, self-report questionnaires, and a 13-minute eyes-open resting-state fMRI scan.

### Interview and self-report measures

#### Clinician Administered PTSD Scale for DSM-5 (CAPS-5)

The CAPS-5 [43], the gold-standard diagnostic interview for PTSD, was administered by doctoral-level clinicians. This interview consists of 30 items designed to assess the onset, duration, and impact of PTSD symptoms, yielding a determination of PTSD diagnosis and symptom severity. The hyperarousal symptom cluster (Criterion E) consists of 6 symptoms, of which 2 – hypervigilance and exaggerated startle response – form an “anxious arousal” symptom factor [44].

#### Life Events Checklist (LEC-5)

The LEC-5 [45] is a 17-item assessment of potentially traumatic events used to determine which events a participant has experienced, witnessed, or learned about happening to a family member or close friend, reflecting a Criterion A trauma.

### PACAP levels

Blood samples were collected at the beginning of each study visit, between approximately 8:00-10:00 AM. Human plasma samples were prepared as described previously [11,46], and all human PACAP38-specific measurements were performed at the University of Vermont, Larner College of Medicine, using double antibody sandwich ELISA immunoassays (Cat. No. HUFI02692, AssayGenie, Dublin, Ireland). We excluded four samples from our analyses due to undetectable PACAP levels. Outliers with exceedingly high (n = 2) or unreliably low (n = 1) concentrations of PACAP levels were winsorized to the next highest or lowest reliable/non-outlier values [46]. Additional details are reported in the Supplemental Methods.

### MRI data acquisition and preprocessing

Imaging was conducted at the McLean Hospital Imaging Center on a 3T Siemens Prisma scanner with a 64-channel head coil. Structural and functional images were initially acquired using the Human Connectome Project (HCP) Young Adult protocols (n = 15). Early in the study, imaging protocols were transitioned to the HCP Lifespan protocols (n = 78). Protocol details are documented in the Supplement. These protocols were developed to be compatible with each other [47]. As such, no effect of scanner on measures of interest was found (Supplemental Results). Results are therefore reported for the entire sample (Young Adult + Lifespan). Lifespan protocol findings are reported in the Supplemental Results.

MRI data were preprocessed using fMRIPrep version 20.2.7 [48], which is based on Nipype 1.7.0 [49]. Participants were excluded if their mean framewise displacement (FD) exceeded 0.5 mm or greater than 20% of volumes exceeded FD = 0.5 mm (n = 7) [50]. Further preprocessing details are presented in the Supplemental Methods.

### Resting-state functional connectivity analyses

Additional preprocessing of resting-state fMRI data was conducted using the CONN toolbox [51], including the regression of physiological noise from white matter and cerebrospinal fluid using the CompCor method [52], scrubbing of motion outliers (FD > 0.5 mm) [50], and high pass (0.01 Hz) filtering.

Cleaned timeseries were extracted from standard left/right (l/r) amygdala (AMYG) regions of interest (ROIs) [53] and DMN ROIs. Pearson correlation analyses were performed between l/r AMYG and DMN ROI timeseries to generate individual pairwise ROI-based AMYG functional connectivity (FC) values. DMN ROIs consisted of medial prefrontal cortex (mPFC), left and right angular gyrus (l/rANG), and combined posterior cingulate cortex-precuneus (PCC-Precun) masks taken from the Brainnetome Atlas, given its superior segregation of cortical subregions based on functional and anatomical connectomics [54]. CeA and BLA AMYG subregion ROIs were generated from the JuBrain Atlas [55,56] – note that the JuBrain Atlas combines both CeA and medial AMYG nuclei into a single centromedial (CMA) ROI, as segmentation of these adjacent subnuclei has been unreliable in human 3T fMRI (**Figure 1)**. Whole-brain seed-based correlations were performed for AMYG seeds to compute whole-brain AMYG FC maps. ROI- and seed-based FC values were Fisher’s z-transformed prior to further statistical analyses.

**Figure 1.**
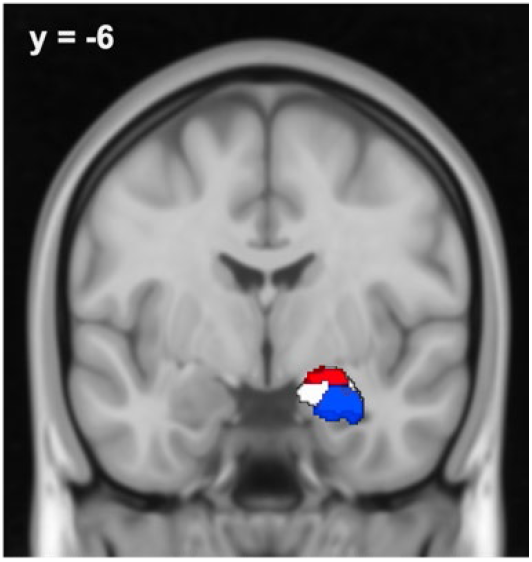
Amygdala subregion ROIs. JuBrain Atlas centromedial (CMA, red) and basolateral (BLA, blue) amygdala subregion ROIs [53] overlaid on the canonical Harvard-Oxford amygdala ROI (white) [50]. Only the right hemisphere is depicted.

### Statistical analyses

Pearson correlation analyses examined associations between circulating PACAP levels and pairwise ROI-based FC of the l/rAMYG with DMN ROIs. Significant effects were followed by whole-brain analyses to demonstrate the robustness and spatial specificity of the effects. Circulating PACAP levels were regressed onto whole-brain connectivity maps seeded in AMYG ROIs, demonstrating significant effects using SPM12. Whole-brain results were thresholded at p < 0.005 (uncorrected height threshold), p < 0.05 FDR-corrected cluster-size threshold. Whole-brain results not surviving correction are reported in the Supplemental Results.

Based on *a priori* interest in sex- and AMYG subregion-specific effects, all analyses were additionally performed on male and female participants separately, as well as CMA and BLA ROIs separately. Correlation coefficients between groups (male vs. female) and subregions (CMA vs. BLA) were compared using Fisher’s and Hittner’s r to z procedure, respectively, through the cocor package in R [57], using one-tailed tests given strong *a priori* hypotheses.

FC effects demonstrating a significant association with PACAP were submitted to Pearson correlation analyses with CAPS-Anxious Arousal symptom severity for clinical association analyses. Sensitivity analyses were performed across CAPS-Hyperarousal symptoms to test the unique association with startle response specifically, given its demonstrated association with the PACAP system.

All analyses were rerun with age, sex, PTSD diagnosis, and total lifetime trauma exposure as covariates, yielding equivalent results (Supplemental Results).

## RESULTS

### Circulating PACAP levels are associated with increased connectivity between the amygdala and posterior DMN

Across the whole sample, ROI-based functional connectivity analyses revealed a positive association between circulating PACAP levels and rAMYG-PCC/Precun connectivity (**Figure 2A**; r = 0.25, p = 0.019, FDR p < 0.05) and rAMYG-lANG connectivity (r = 0.30, p = 0.004, FDR p < 0.05), but not rAMYG-mPFC (r = -0.03, p = 0.756) or rAMYG-rANG (r = 0.15, p = 0.162). No effects were seen with the lAMYG (p’s > 0.298).

**Figure 2.**
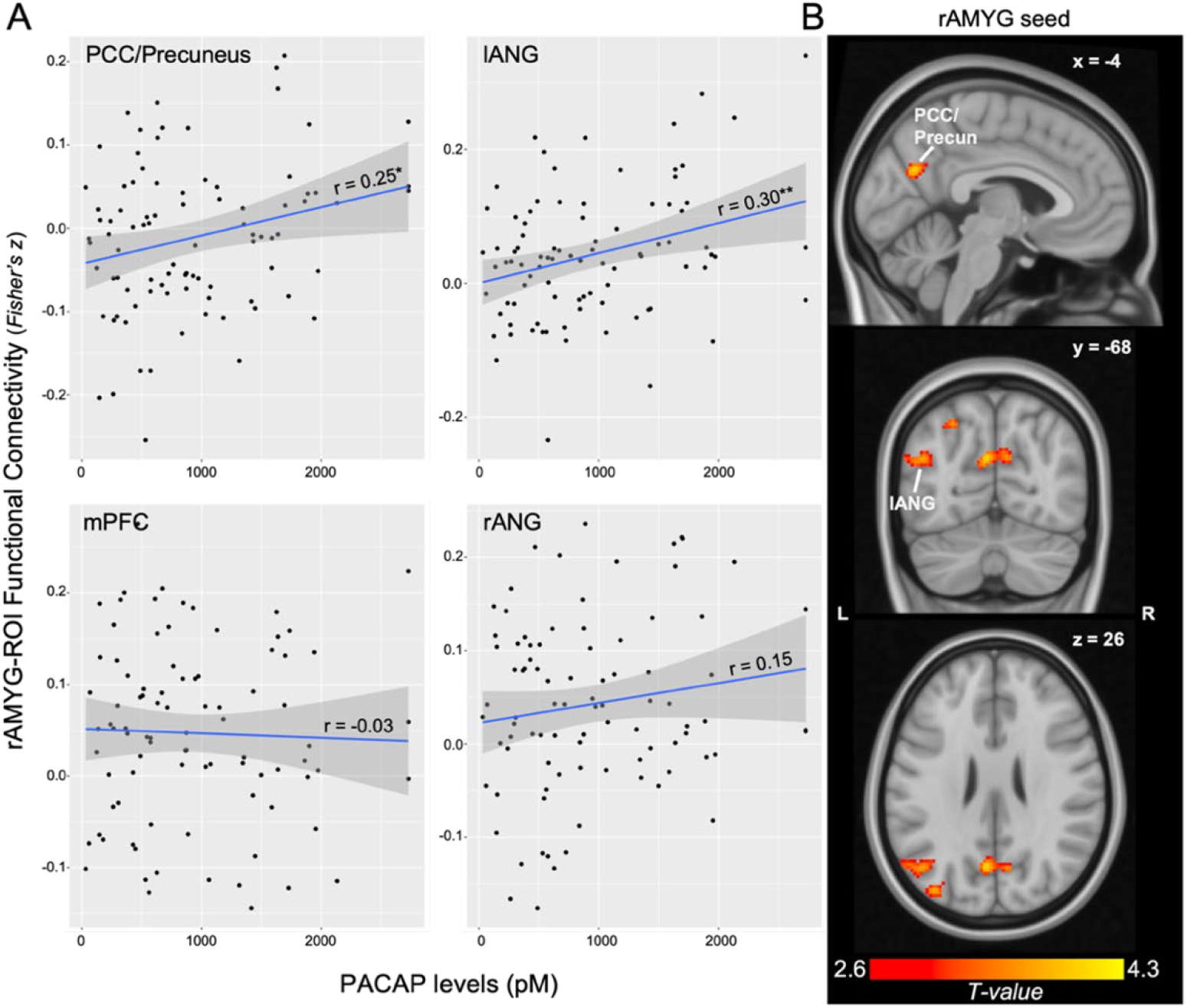
PACAP and AMYG connectivity. A) Scatterplot of ROI-based rAMYG-DMN connectivity with PACAP levels, demonstrating a unique association with posterior DMN nodes. B) Whole-brain regression of PACAP levels on rAMYG seed connectivity, demonstrating the spatial specificity of these effects. Display threshold set at voxel-wise p < 0.005, k = 50. PCC = posterior cingulate cortex; l/rANG = left/right angular gyrus; mPFC = medial prefrontal cortex. \**p* < 0.05, ** *p* < 0.01.

Whole-brain regression of PACAP levels on rAMYG-seed connectivity confirmed the spatial specificity of these effects. Circulating PACAP levels were associated with greater connectivity between the rAMYG and a cluster spanning the bilateral PCC/Precuneus (**Figure 2B**; k = 278, cluster FDR q = 0.006, peak = -4, -68, 26, T = 4.29) and a cluster in the lANG (k = 245, cluster FDR q = 0.006, peak = -38, -82, 26, T = 3.83). No other significant clusters emerged, and no negative associations were seen.

#### Sex-specific analyses

Planned analyses within each sex revealed that PACAP associations with rAMYG-PCC/Precun and rAMYG-lANG connectivity were statistically significant only in female participants (**Figure 3A**; PCC/Precun: r = 0.29, p = 0.016, FDR p < 0.05; lANG: r = 0.34, p = 0.005, FDR p < 0.05). Correlations in the male participants were not significant and corresponded to smaller effect sizes (PCC/Precun: r = -0.18, p = 0.452; lANG: r = 0.06, p = 0.808). Comparison of the correlation coefficients identified a significantly stronger association in females than males for rAMYG-PCC/Precun (z = 1.77, p = 0.039 one-tailed) but not rAMYG-lANG connectivity (z = 1.08, p = 0.140 one-tailed).

**Figure 3.**
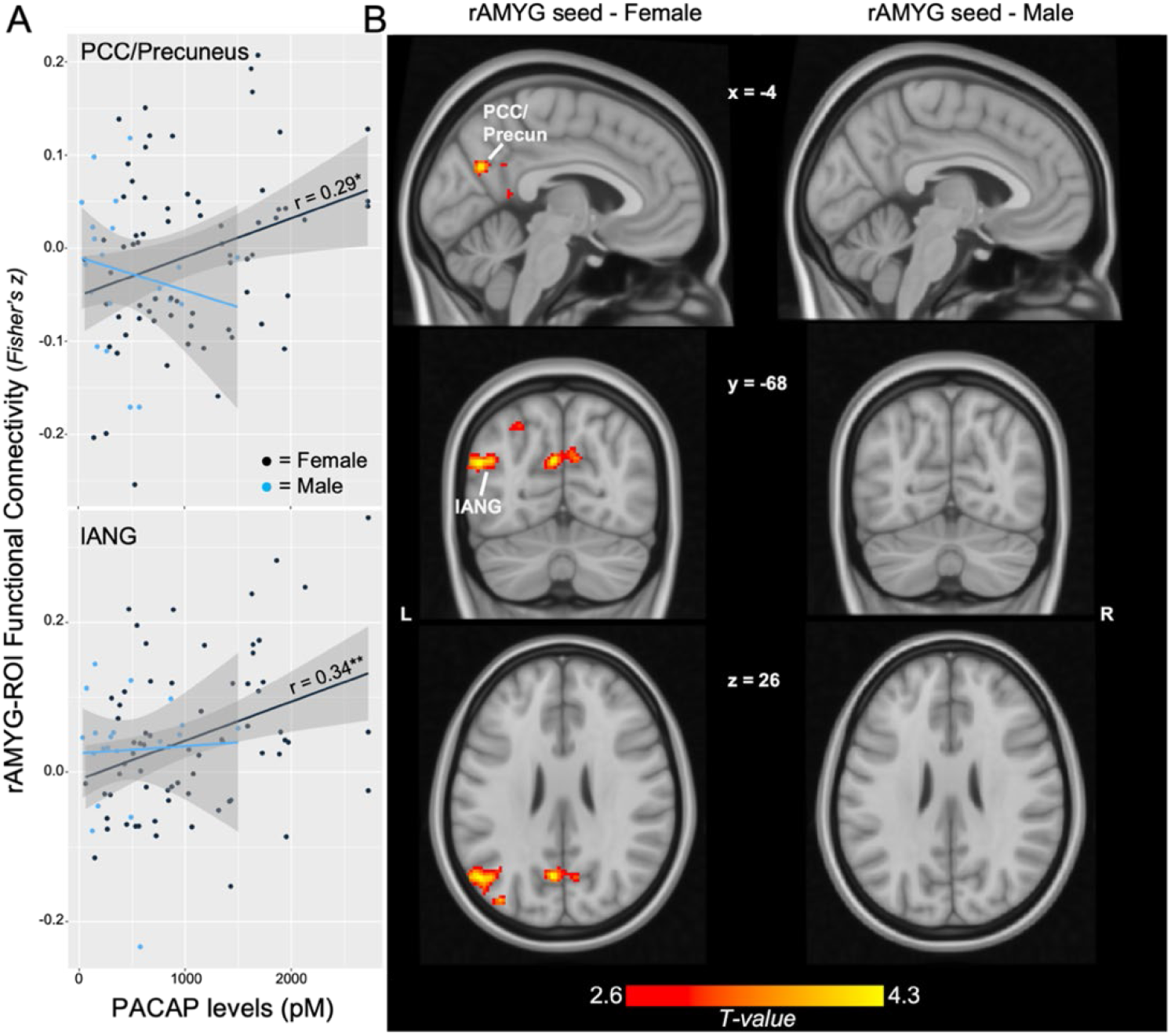
PACAP and AMYG connectivity in female and male participants. A) Scatterplot of ROI-based rAMYG-PCC/Precun connectivity with PACAP levels for female vs. male participants, demonstrating effects only in females. B) Female whole-brain regression of PACAP on rAMYG seed connectivity demonstrated the spatial specificity of these effects. Display threshold set at voxel-wise p < 0.005 uncorrected, k > 50. Depicted coordinates are those of the main rAMYG effects in the whole sample, rather than those of the female-only participants, to illustrate similarities in cluster location and extent across analyses given the use of cluster-based statistics. \**p* < 0.05, ** *p* < 0.01.

Whole-brain regression of PACAP levels on rAMYG-seed connectivity in female participants confirmed this effect (**Figure 3B**; PCC/Precun: k = 319, cluster FDR q = 0.001, peak = 12, -54, 32, T = 4.09; lANG: k = 296, cluster FDR q = 0.001, peak = -52, -68, 24, T = 4.15). No other significant clusters emerged.

#### Amygdala subregion analyses

Across the whole sample, PACAP showed a non-significant trend association with CMA-PCC/Precun connectivity (r = 0.20, p = 0.057) but not BLA-PCC/Precun connectivity (r = 0.09, p = 0.363). Comparison of correlation coefficients revealed no significant differences in strength of association with PACAP between the CMA and BLA (p = 0.159 one-tailed). PACAP also was not significantly associated with connectivity of AMYG subregions with the lANG (CMA r = 0.17, p = 0.101; BLA r = 0.18, p = 0.088). Whole-brain analyses revealed similar patterns - the rCMA demonstrated a positive association between circulating PACAP levels and connectivity with the PCC/Precun (k = 193, cluster FDR q = 0.030, peak = 10, -56, 32, T = 4.11) but not with the lANG. No whole-brain effects emerged with the rBLA ROI.

Within the female participants, PACAP was significantly associated with CMA-PCC/Precun connectivity (r = 0.25, p = 0.036; **Figure 4A**), but not with BLA-PCC/Precun connectivity (r = 0.15, p = 0.223). There were no significant differences in strength of correlation between the CMA and BLA (p = 0.196 one-tailed). Within males, neither of these associations was statistically significant (CMA: r = -0.06, p = 0.795; BLA: r = -0.34, p = 0.147). Whole-brain analyses confirmed the spatial specificity of this CMA-effect in females to a bilateral PCC/Precun cluster (**Figure 4B**, k = 201, cluster FDR q = 0.021, peak = 10, -54, 34, T = 4.06), with no such effects emerging with the BLA.

**Figure 4.**
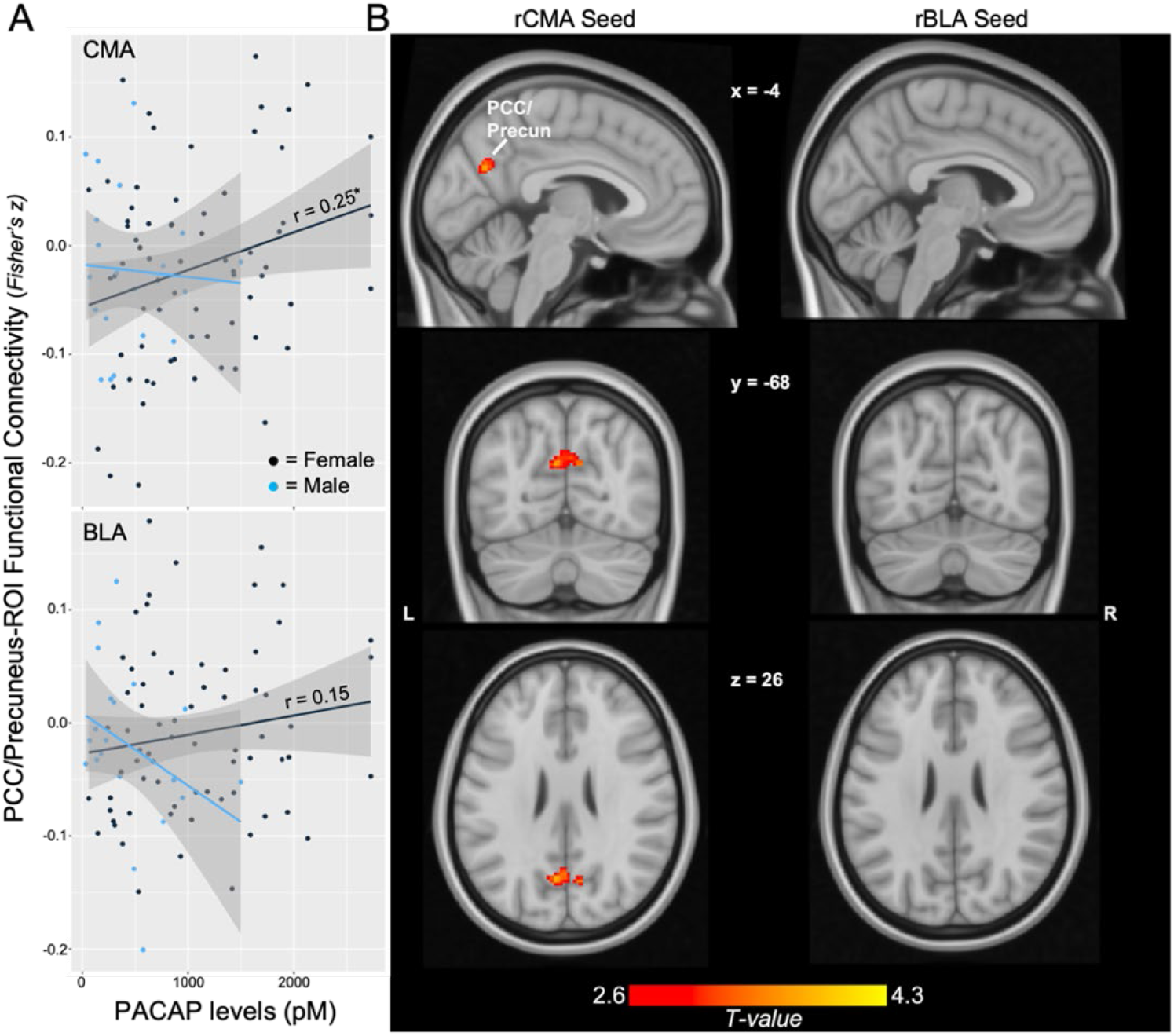
AMYG subregion effects. A) Scatterplot of CMA- and BLA-PCC/Precun connectivity with PACAP levels for females vs. males. B) Female whole-brain regression of PACAP on rCMA, rBLA seed connectivity. Display threshold set at voxel-wise p < 0.005 uncorrected, k > 50. Depicted coordinates mirror those of the main rAMYG effects, and not CMA-specific peak voxels, to demonstrate similarities in cluster location and extent across analyses given the use of cluster-based statistics. \**p* < 0.05, ** *p* < 0.01.

### Clinical Associations

Clinical correlation analyses revealed that rAMYG-PCC/Precun connectivity was significantly associated with CAPS-Anxious Arousal scores (**Figure 5A**, r = 0.23, p = 0.025). Sensitivity analyses revealed a specific association of this connectivity with the CAPS – Startle Response (r = 0.23, p = 0.025) and no other hyperarousal symptoms (p’s > 0.142). This effect was not sex-specific (females/males r = 0.20/0.36, p = 0.096/0.100). No clinical associations were seen for rAMYG-lANG connectivity (p’s > 0.273). Evaluation of AMYG subregions revealed no unique subregion effects (p’s > 0.128).

**Figure 5.**
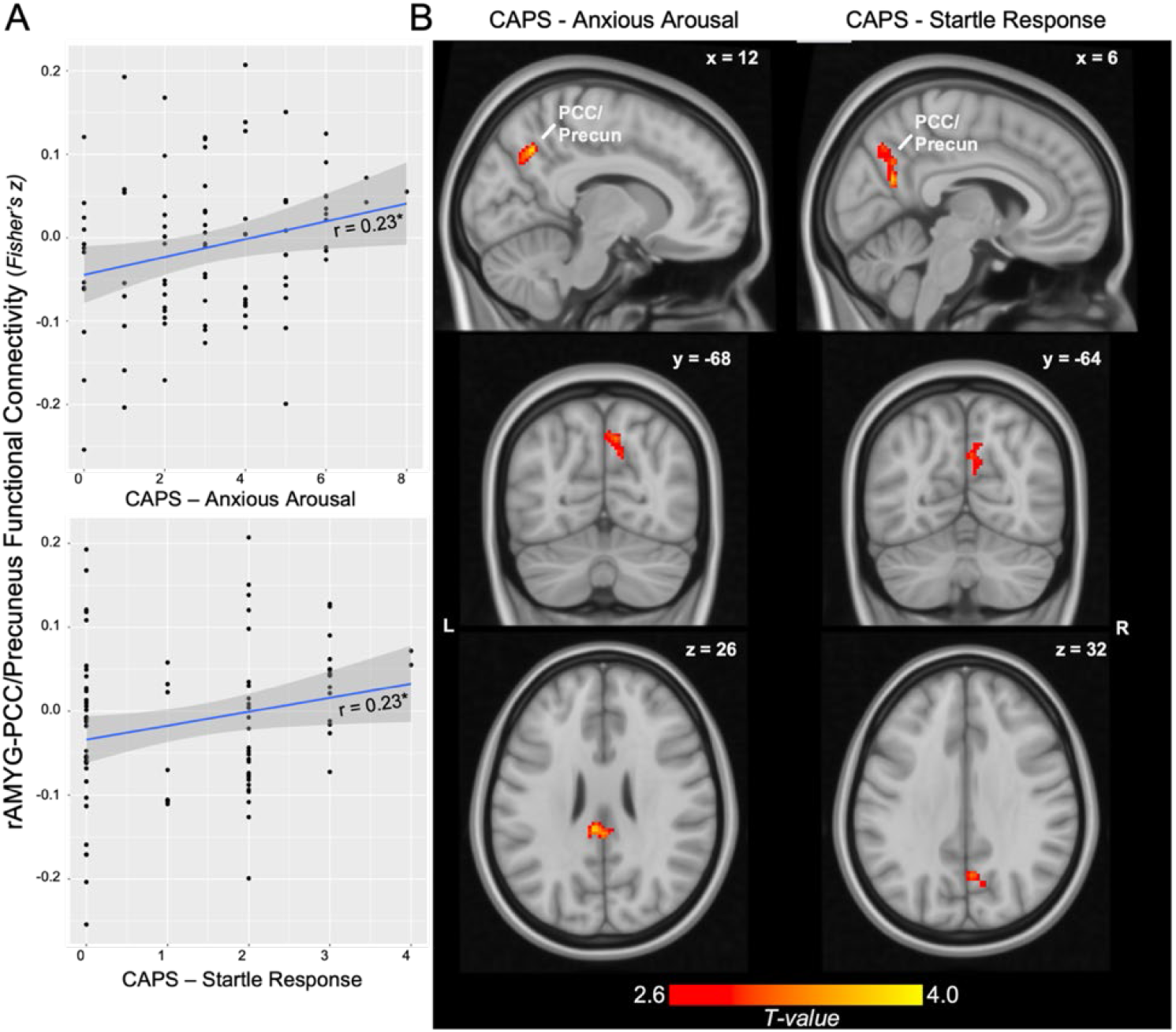
Clinical associations of rAMYG-PCC/Precun connectivity with Anxious Arousal symptoms. A) Scatterplot of rAMYG-PCC/Precun connectivity with CAPS-Anxious Arousal and Startle Response symptom scores. B) Whole-brain regression of CAPS-Anxious Arousal and Startle Response symptom scores on rAMYG seed connectivity. Display threshold set at voxel-wise p < 0.005 uncorrected, k > 50.

Whole-brain regression of CAPS-Anxious Arousal demonstrated a positive association with increased connectivity between rAMYG and a cluster spanning the right PCC/Precuneus (**Figure 5B**, k = 158, cluster FDR q = 0.082, peak = 12, -60, 40, T = 3.78) as well as a bilateral cluster in the dorsal PCC (k = 138, cluster FDR q = 0.082, peak = -6, -26, 32, T = 3.93). CAPS-Startle Response was associated with increased connectivity between the rAMYG and a cluster spanning the right PCC/Precuneus; however, this did not survive correction (k = 122, cluster FDR q = 0.166, peak = 6, -62, 22, T = 3.56).

## DISCUSSION

Expanding upon the growing literature demonstrating an influence of PACAP on amygdala activity, we demonstrate here that circulating levels of PACAP are associated with increased intrinsic functional connectivity between the amygdala and posterior regions of the DMN. A similar connectivity profile is associated with anxious arousal symptoms, specifically startle response, which is consistent with prior work demonstrating a unique effect of PACAP on arousal and threat reflexes. In keeping with this prior work, our data suggest these PACAP effects on amygdala connectivity may be unique to females and, to a lesser extent, the centromedial amygdala; however, these effects should be interpreted with caution (see below).

These findings emerge in the context of accruing evidence for exaggerated coupling between the intrinsically anti-correlated SN and DMN in PTSD. Exaggerated connectivity between the amygdala (SN) and the PCC/Precuneus (DMN) has been implicated in risk, severity, and maintenance of PTSD [27,31,58,59]. Such connectivity patterns are similarly implicated in transdiagnostic processes of aberrant stress response [29,33], threat reactivity [32], and trait anxiety [60,61]. Functionally, this specific circuit, and broader SN-DMN coupling, is suggested to regulate attentional deployment to salient threat cues, with intrinsic anti-correlation between these structures/networks reflecting homeostatic inhibition of threat orientation at rest [26,28,62,63]. Our clinical association analyses support this notion, with increased AMYG-PCC/Precun connectivity demonstrating unique associations with subjective reports of exaggerated startle response. This key translational threat-orienting behavior is known to be affected by the PACAP system [25,64]. Taken together, these findings align with the proposed role of PACAP signaling within the amygdala to elicit stress responses and behavioral reflexes to threat.

This purported association of PACAP with behavioral threat reflexes is further supported by the specificity of our PACAP findings to the right amygdala and, to a lesser extent, the CMA. Post-hoc analyses revealed a significant effect of hemisphere (left vs. right AMYG) on the association between PACAP and AMYG connectivity (p = 0.013). Although findings regarding laterality of amygdala structure and function are heterogeneous, a recent review of amygdala laterality suggests a right-hemispheric dominance in neuropeptide expression, specifically under stress, and functional specialization in rapid fear/threat processing and responding [65]. However, the authors note lateralization of subnuclei effects remains understudied in humans, due in part to limits to the spatial resolution of human functional imaging. Notably, our analyses utilized a combined centromedial ROI, which may have contributed to the lack of significant contrasts of CMA vs. BLA connectivity, as evidence for PACAP influence on medial amygdala activity is sparse [66]. The lack of significant differences between the association of CMA and BLA connectivity with PACAP precludes any statements of subregion specificity from the current sample. Nonetheless, that the effects emerged in the CMA and not the BLA provides preliminary evidence for potential subregion differences. Therefore, future studies utilizing more refined parcellations of amygdala nuclei, including the adjacent BNST, are needed to ascertain subnuclei specificity of PACAP effects in humans. Keeping these considerations in mind, our findings align with the extant literature suggesting an effect of PACAP on behavioral threat responses via central amygdala signaling [20,25,67]. That these effects are present even at rest suggests a “rewiring” of the intrinsic functional architecture of the amygdala by PACAP to prime behavioral responses to threat.

While the cross-sectional nature of our data limits the determination of causal mechanisms linking PACAP with aberrant intrinsic amygdala connectivity, answers may lie in the demonstrated neuroplastic effects of PACAP. Rodent studies have identified robust neurotrophic [68,69] and neuroplastic [70–72] properties of PACAP, especially within the central extended amygdala [73], which are enhanced under stress [21]. Such properties parallel general stress-induced increases in neuroplasticity within the central extended amygdala [74,75], which has been linked to sustained anxiety-like states through strengthened synaptic transmission between neural structures recruited in threat processing [76,77]. Notably, acute stress results in activation and increased coupling of central SN and DMN hubs [30]. Sustained effects of stress are further associated with co-activations of the amygdala and posterior midline DMN structures, including the PCC/Precuneus [29,33], which together demonstrate similar metabolic profiles in response to reactivation of threat memory [78]. As such, the enhanced neuroplastic properties of PACAP under stress may strengthen the synaptic transmission between the co-activated amygdala and PCC/Precuneus, resulting in lasting increases in intrinsic functional connectivity and sustained changes in resting-state network dynamics. Investigating the directionality and causality of PACAP signaling could lead to promising therapeutic avenues for modulating its effects through the right combination of agonism and antagonism of its receptors [79].

Associations between PACAP and amygdala connectivity emerged within the angular gyrus (ANG) as well. Positioned as a lateral node of the DMN, the ANG is a densely connected structure that sits at the junction of multiple functional systems and has thus been implicated as a cross-modal hub [80]. Less frequently implicated in states of stress or threat-mediated arousal, the observed functional coupling between the amygdala and lANG may reflect broader increases in amygdala-DMN coupling. Additionally, as a cross-modal hub linking external sensory processing with internal, self-referential representations and memories [81,82], this amygdala-lANG coupling may reflect higher-order threat processes, such as threat-associated memory retrieval. This is supported by a lack of association between PACAP and lANG connectivity with the CMA or with the lower-order startle response symptom, suggesting a general coupling with the amygdala that is not specific to threat-reactive reflexes.

Surprisingly, we found no associations between PACAP levels and amygdala-prefrontal cortex connectivity. The amygdala-prefrontal circuit has been at the forefront of neurocircuitry models of PTSD for decades [15,16,83–85] and has been reliably implicated in the regulation of fear acquisition, expression, and extinction [86–88]. Evidence suggests PACAP is markedly expressed within prefrontal (infralimbic in rodents) structures, through which it may play a mechanistic role in fear learning and extinction [89]. However, prior human studies have similarly found no association between the PACAP-PAC1R system and prefrontal activity in relation to PTSD or threat reactivity [19]. Additionally, structures of the medial prefrontal cortex serve as anterior hubs of the DMN which are disrupted in PTSD [90]; however, our findings demonstrate a unique effect with posterior DMN hubs. Notably, the PCC/Precuneus and amygdala are positioned at the apex of the cortical and subcortical functional connectivity hierarchy, respectively [91,92], and may thus serve as intrinsic connectivity hubs. Therefore, PACAP’s effects on intrinsic connectivity of the amygdala may be unique to these resting-state hubs, whereas its influence on the prefrontal cortex may be task-dependent. Moreover, heterogeneity in prefrontal cortex findings in PTSD [93,94] mirrors the heterogeneity of prefrontal subregions and their distinct task-dependent functions [86,95]. More comprehensive investigations into the role of PACAP in the intrinsic connectivity of the human PFC are needed to elucidate these spatial and functional nuances.

Our data suggest that the purported role of PACAP on intrinsic amygdala connectivity may be sex specific. Direct comparisons between sexes in the current sample should be interpreted with caution given significant discrepancies in sample sizes. Keeping this limitation in mind, our contrasts of correlation coefficients revealed stronger associations in females. This aligns with initial work demonstrating an association between circulating PACAP levels and PTSD that is unique to female patients [11], as well as preclinical work demonstrating estrogenic modulation of the PACAP-PAC1R system [11,42,96]. The observed sex differences in PACAP levels in our sample (**Table 1**) were robust to other demographic and clinical variables, lending further credence to the potential sexual dimorphism of the PACAP system in PTSD (Supplemental Results). Moreover, the identified PACAP-associated AMYG-PCC/Precun circuit has been previously shown to be sex-dependent and uniquely elevated in female patients with PTSD [34]. Therefore, PACAP-associated alterations of intrinsic amygdala connectivity may yield important insights into our mechanistic understanding of the increased prevalence and risk of PTSD in individuals assigned female at birth. Future studies matching biological sex groups and investigating gonadal hormone levels are needed to delineate the true sex-specificity of PACAP’s effects on intrinsic amygdala connectivity in humans.

Finally, as the current study investigated circulating serum PACAP levels, it is unclear if this association between PACAP and amygdala-DMN connectivity reflects a risk marker for PTSD, and/or physiological sequalae of the condition. Genetic variants associated with an altered PACAP-PAC1R system have been implicated as risk markers for PTSD through changes in neural activity [11,97]. Moreover, genetic risk for related dysfunction of the HPA axis and stress-sensitive serotonergic systems has been linked to amygdala and PCC/Precuneus activity [98–100]. Additionally, recent work in non-PTSD anxious patients also supports a PACAP-related relationship to anxiety- or stress-sensitivity [46]. Taken together, these data position this circuit and its association with PACAP as a potential intermediate phenotype of interest. However, phasic elevations of circulating PACAP also have been found after acute and sustained stress, reflecting a state-like process or prolonged stress-response [25,101], and mirror the state-like alterations of amygdala-PCC/Precuneus connectivity in response to acute and chronic stress. Therefore, evaluations of PACAP-PAC1R risk genes and pathways are needed to further elucidate the role of PACAP-associated intrinsic amygdala connectivity in risk or pathophysiology models of PTSD.

The present findings provide initial evidence for an association between PACAP and intrinsic amygdala functional connectivity in PTSD. That the identified network has been reliably implicated in PTSD risk and symptom severity, as well as related transdiagnostic processes, adds to the growing body of evidence implicating the PACAP system in PTSD and stress-related arousal. Moreover, it offers novel mechanistic insights into dysfunctional neural circuitry in PTSD and how the PACAP system may confer risk for PTSD through disruption of intrinsic resting-state network dynamics.

## Data Availability

Data produced in the present study are available upon reasonable request to the authors

## Acknowledgements

This work was supported by NIH awards P50-MH115874 (to WAC, KJR; Project 4: IMR, SLR), R01-MH120400 (IMR), and R01-MH97988 (SEH and VM).

## SUPPLEMENTAL MATERIALS

### MATERIALS AND METHODS

#### Participants

Inclusion criteria included ability to provide written informed consent, 18-55 years old, and any gender. Given our interest in sex differences, specifically the influence of estrogen modulation of PACAP, participants were required to be the same sex as assigned at birth, female subjects were to be premenopausal, and participants with a history of receiving hormonal replacement therapy or undergoing surgery to change biological sex were excluded. Other exclusion criteria included left-handedness, medical conditions that would confound results, such as a seizure or other neurological disorder, inability to tolerate blood draws, history of moderate to severe traumatic brain injury, current treatment with an antipsychotic (unless prescribed only for sleep), MR contraindications, including metal implants and claustrophobia, positive pregnancy test for female participants on the day of scanning. In addition, participants were excluded for lifetime history of schizophrenia or schizoaffective disorder and if they met for current (past month) moderate-to-severe alcohol or substance use disorder, psychotic disorder, anorexia, obsessive compulsive disorder, or manic or mixed mood episode.

#### PACAP Assays

Participants were instructed not to eat the morning of their visit prior to their blood draw. Samples were centrifuged at 3500 rpm for 15 minutes. Plasma was extracted and stored at -80 °C until analysis. Optimal sample volume was determined in dilution tests, and all values represent the mean from assay duplicates; intra-assay variation was approximately 9%. Assay midpoint was 1.1 fmol and detection limit from the linear range of the standard curve was 0.2 fmol.

#### MRI data acquisition and preprocessing

T1-weighted 3D MPRAGE structural images were initially acquired using the HCP 0.7mm resolution sequence (TR/TE: 2400/2.14 ms; flip angle: 8 deg; FOV: 224 × 224; voxel size: 0.7mm isotropic), and eyes-open resting state T2-weighted echoplanar images were initially acquired using the HCP Young Adult sequence (TR/TE: 720/33.1 ms, in-plane resolution: 2mm; voxels: 2mm isotropic; multiband factor = 8; two runs of 600 frames each). Early in the study, imaging protocols were transitioned to the HCP Lifespan protocol (n = 78) – T1-weighted 3D MPRAGE structural images were acquired using the HCP 0.8mm resolution sequence (TR/TEs: 2500/1.81/3.6/5.39/7.18; flip angle: 8 deg; FOV: 256 × 240; voxel size: 0.8mm isotropic), and eyes-open resting state T2-weighted echoplanar images were acquired using the HCP Lifespan sequence (TR/TE: 800/37 ms, in-plane resolution: 2mm; voxels: 2mm isotropic; multiband factor = 8; anterior-posterior phase encoding; two runs of 488 frames each).

T1-weighted (T1w) images were corrected for intensity non-uniformity [1]. Brain tissue segmentation of cerebrospinal fluid (CSF), white matter (WM) and gray matter (GM) was performed on the brain-extracted T1w [2]. Brain surfaces were reconstructed using recon-all [3], and the brain mask estimated previously was refined with a custom variation of the method to reconcile ANTs-derived and FreeSurfer-derived segmentations of the cortical GM of Mindboggle [4]. Volume-based spatial normalization to MNI standard space (MNI152NLin6Asym) was performed through nonlinear registration with antsRegistration (ANTs 2.3.3), using brain-extracted versions of both T1w reference and the T1w template.

EPI images were corrected for susceptibility distortions using the fMRIPrep fieldmap-less approach [5]. Based on the estimated susceptibility distortion, a corrected EPI (echo-planar imaging) reference was calculated for a more accurate co-registration with the anatomical reference. The reference was co-registered to the T1w reference with six degrees of freedom [6]. Head motion parameters with respect to the reference (transformation matrices, and six corresponding rotation and translation parameters) were estimated before any spatiotemporal filtering [7]. EPI images were slice-time corrected [8]. The time series were resampled onto their original, native space by applying a single, composite transform to correct for head motion and susceptibility distortions. The time series were resampled into standard space, generating a preprocessed run in MNI152NLin6Asym space. First, a reference volume and its skull-stripped version were generated using a custom methodology of fMRIPrep. Automatic removal of motion artifacts using independent component analysis (ICA-AROMA) [9] was performed on the preprocessed images on MNI space time-series after removal of non-steady state volumes and spatial smoothing with an isotropic, Gaussian kernel of 6mm FWHM (full-width half-maximum). Corresponding “non-aggressively” denoised runs were produced after such smoothing [9].

### RESULTS

#### Sex-differences in circulating PACAP levels

Female participants demonstrated significantly greater circulating PACAP levels than males (t = 5.34, p < 0.001). These differences were robust to controlling for other clinical and demographic variables demonstrating sex differences (age and total PTSD symptom severity; F = 14.60, p < 0.001), suggesting sex-related differences in circulating PACAP levels were not attributable to other differences.

#### No effect of scanner protocol

The results were robust to controlling for scanner protocol. Circulating PACAP levels were associated with rAMYG-PCC/Precun (semi-partial r = 0.25, p = 0.011) and rAMYG-lANG connectivity (sr = 0.30, p = 0.004). Sex-specific effects similarly held – in women, circulating PACAP levels were associated with rAMYG-PCC/Precun (sr = 0.30, p = 0.014) and rAMYG-lANG connectivity (sr = 0.34, p = 0.005). No effects emerged for men (p’s > 0.364). Scanning protocols did not differ in any of the AMYG FC values (p’s > 0.268).

#### Lifespan protocol effects

ROI-based functional connectivity analyses revealed a positive association between circulating PACAP levels and rAMYG-PCC/Precun connectivity (r = 0.30, p = 0.009, FDR p < 0.05) and rAMYG-lANG connectivity (r = 0.36, p = 0.002, FDR p < 0.05), but not rAMYG-mPFC (r = 0.03, p = 0.796) or rAMYG-rANG (r = 0.19, p = 0.099). No effects were seen with the lAMYG (p’s > 0.298).

Planned analyses of sex-specific effects revealed PACAP associations with rAMYG-PCC/Precun and rAMYG-lANG connectivity were present only in female participants (PCC/Precun: r = 0.38, p = 0.004, FDR p < 0.05; lANG: r = 0.40, p = 0.002, FDR p < 0.05). No associations were seen in male participants (PCC/Precun: r = -0.22, p = 0.378; lANG: r = 0.06 p = 0.807).

Planned analyses of AMYG subregion-specific effects suggested associations with PACAP were somewhat unique to the CMA. PACAP was associated with CMA-PCC/Precun connectivity (r = 0.25, p = 0.031) but not BLA-PCC/Precun connectivity (r = 0.16, p = 0.171). No such specificity was seen for the lANG (CMA r = 0.22, p = 0.058; BLA r = 0.23, p = 0.052).

Subregion effects with PCC/Precun connectivity were similarly specific to females (CMA: r = 0.34, p = 0.010; BLA: r = 0.24, p = 0.078) and not males (CMA: r = -0.11, p = 0.670; BLA: r = -0.33, p = 0.172). Whole-brain analyses again confirmed the spatial specificity of this CMA-effect in females to a bilateral PCC/Precun cluster (**Figure 4B**, k = 201, cluster FDR q = 0.021, peak = 10, -54, 34, T = 4.06), with no such effects emerging with the BLA.

#### Controlling for demographic and clinical variables

The results were robust to controlling for age, sex, PTSD diagnosis, and total life-time trauma exposure (LEC-5) as covariates (sr = semi-partial r). Circulating PACAP levels were associated with rAMYG-PCC/Precun (sr = 0.25, p = 0.023) and rAMYG-lANG connectivity (sr = 0.30, p = 0.005). Sex-specific effects were similarly robust to age, PTSD diagnosis, and total trauma exposure – in women, circulating PACAP levels were associated with rAMYG-PCC/Precun (sr = 0.32, p = 0.008) and rAMYG-lANG connectivity (sr = 0.35, p = 0.004), while no effects emerged for men (p’s > 0.478).

#### Whole-brain results

Whole-brain associations with circulating PACAP levels, including clusters that did not survive correction for multiple comparisons, are provided for the rAMYG seed for the full sample (Table S1) and female participants (Table S2), as well as for the rCMA (Table S3) and rBLA seeds (Table S4).

**Table S1.**
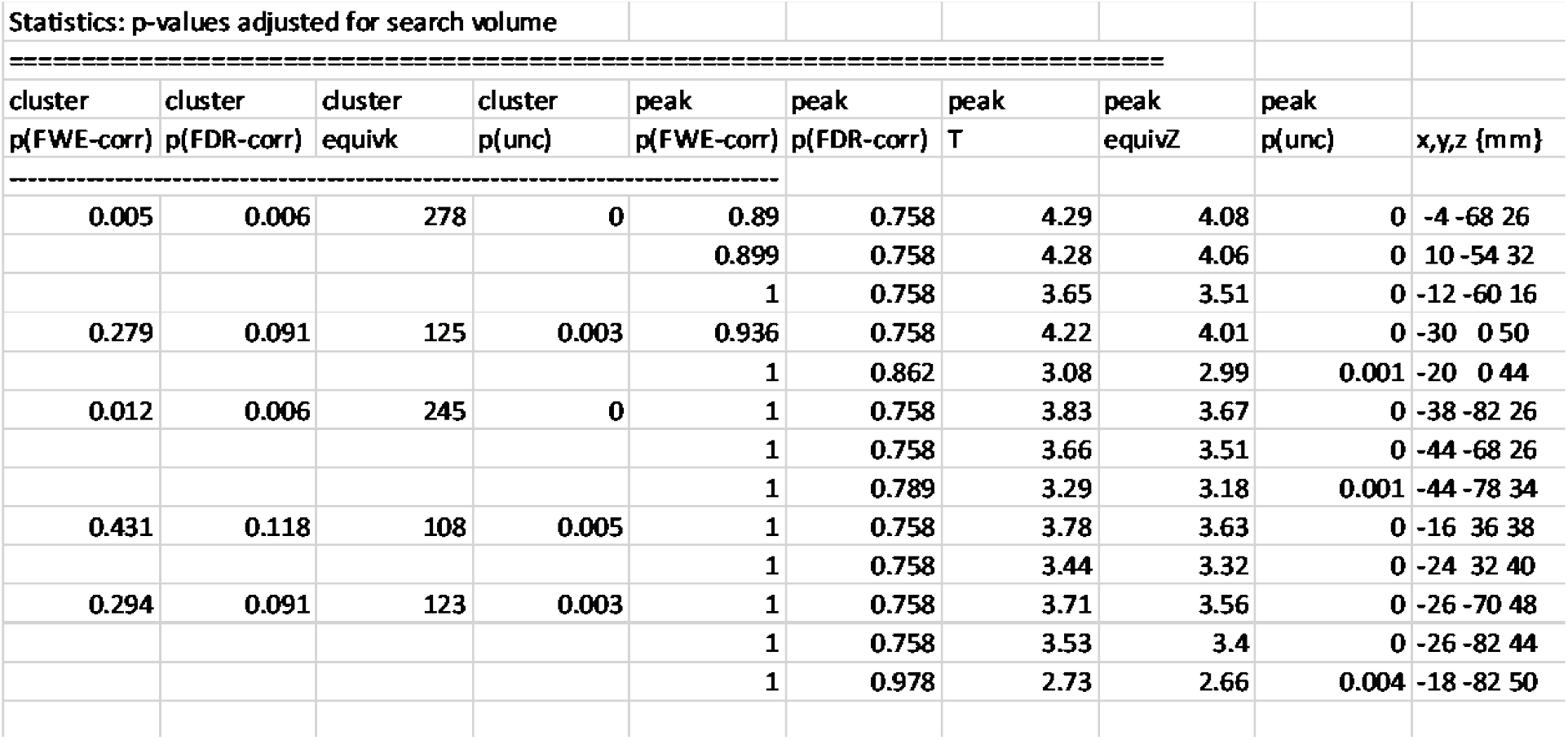
Whole-brain rAMYG seed-based connectivity with circulating PACAP levels. p < 0.005 (uncorrected height threshold), k = 50 arbitrary cluster threshold.

**Table S2.** Whole-brain rAMYG seed-based connectivity with circulating PACAP levels in female participants. p < 0.005 (uncorrected height threshold), k = 50 arbitrary cluster threshold.

| Statistics: p-values adjusted for search volume |  |  |  |  |  |  |  |  |  |
| --- | --- | --- | --- | --- | --- | --- | --- | --- | --- |
| ===== |  |  |  |  |  |  |  |  |  |
| cluster<br>p(FWE-corr) | cluster<br>p(FDR-corr) | cluster<br>equivk | cluster<br>p(unc) | peak<br>p(FWE-corr) | peak<br>p(FDR-corr) | peak<br>T | peak<br>equivZ | peak<br>p(unc) | x,y,z {mm} |
| 0.719 | 0.356 | 81 | 0.01 | 0.861 | 0.994 | 4.43 | 4.13 | 0 | -26 4 46 |
| 0.002 | 0.001 | 296 | 0 | 0.986 | 0.994 | 4.15 | 3.9 | 0 | -52 -68 24 |
|  |  |  |  | 0.993 | 0.994 | 4.1 | 3.86 | 0 | -40 -82 32 |
|  |  |  |  | 1 | 0.994 | 3.35 | 3.21 | 0.001 | -38 -62 24 |
| 0.001 | 0.001 | 319 | 0 | 0.994 | 0.994 | 4.09 | 3.85 | 0 | 12 -54 32 |
|  |  |  |  | 0.996 | 0.994 | 4.06 | 3.82 | 0 | -6 -66 26 |
|  |  |  |  | 1 | 0.994 | 3.83 | 3.63 | 0 | -10 -58 18 |
| 0.809 | 0.358 | 74 | 0.014 | 1 | 0.994 | 3.41 | 3.26 | 0.001 | 34 4 60 |
|  |  |  |  | 1 | 0.994 | 3.29 | 3.16 | 0.001 | 30 2 50 |
| 0.732 | 0.356 | 80 | 0.011 | 1 | 0.994 | 3.29 | 3.16 | 0.001 | -26 -72 48 |
|  |  |  |  | 1 | 0.994 | 3.29 | 3.16 | 0.001 | -26 -82 44 |
|  |  |  |  | 1 | 0.994 | 3.06 | 2.95 | 0.002 | -28 -62 50 |

**Table S3.**
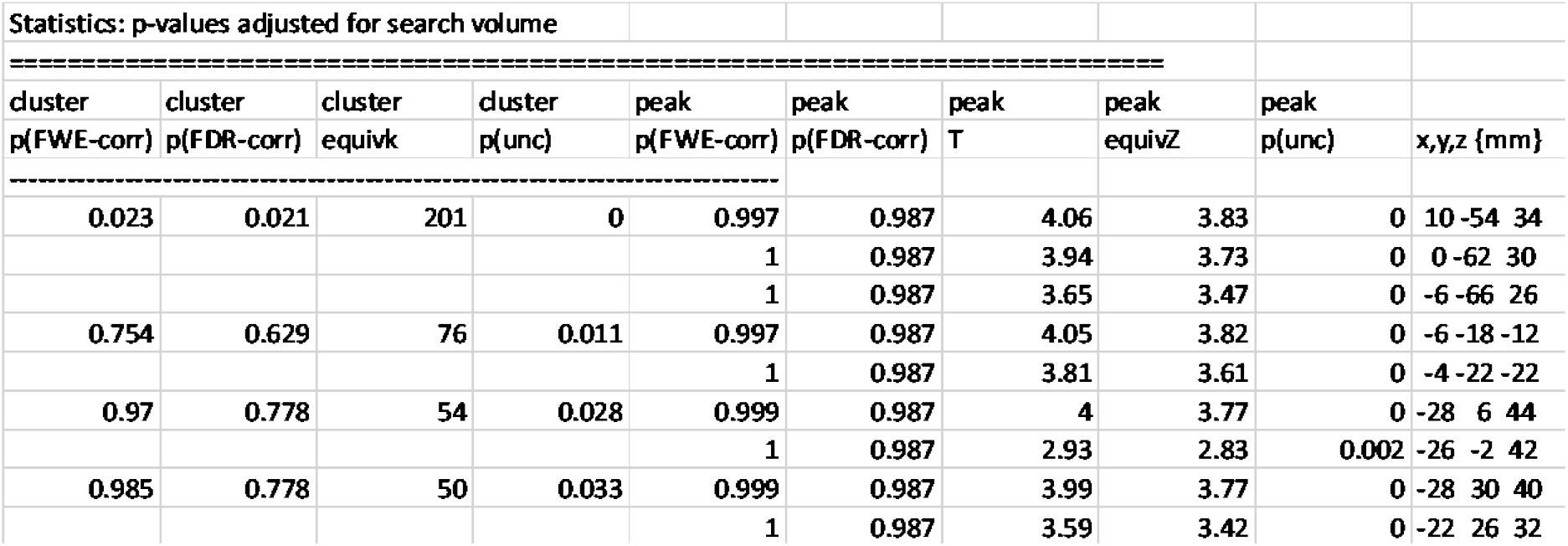
Whole-brain rCMA seed-based connectivity with circulating PACAP levels in female participants. p < 0.005 (uncorrected height threshold), k = 50 arbitrary cluster threshold.

| Statistics: p-values adjusted for search volume |  |  |  |  |  |  |  |  |  |
| --- | --- | --- | --- | --- | --- | --- | --- | --- | --- |
| cluster | cluster | cluster | cluster | peak | peak | peak | peak | peak |  |
| p(FWE-corr) | p(FDR-corr) | equivk | p(unc) | p(FWE-corr) | p(FDR-corr) | T | equivZ | p(unc) | x,y,z {mm} |
| 0.023 | 0.021 | 201 | 0 | 0.997 | 0.987 | 4.06 | 3.83 | 0 | 10 -54 34 |
|  |  |  |  | 1 | 0.987 | 3.94 | 3.73 | 0 | 0 -62 30 |
|  |  |  |  | 1 | 0.987 | 3.65 | 3.47 | 0 | -6 -66 26 |
| 0.754 | 0.629 | 76 | 0.011 | 0.997 | 0.987 | 4.05 | 3.82 | 0 | -6 -18 -12 |
|  |  |  |  | 1 | 0.987 | 3.81 | 3.61 | 0 | -4 -22 -22 |
| 0.97 | 0.778 | 54 | 0.028 | 0.999 | 0.987 | 4 | 3.77 | 0 | -28 6 44 |
|  |  |  |  | 1 | 0.987 | 2.93 | 2.83 | 0.002 | -26 -2 42 |
| 0.985 | 0.778 | 50 | 0.033 | 0.999 | 0.987 | 3.99 | 3.77 | 0 | -28 30 40 |
|  |  |  |  | 1 | 0.987 | 3.59 | 3.42 | 0 | -22 26 32 |

**Table S4.** Whole-brain rBLA seed-based connectivity with circulating PACAP levels in female participants. p < 0.005 (uncorrected height threshold), k = 50 arbitrary cluster threshold.

| Statistics: p-values adjusted for search volume |  |  |  |  |  |  |  |  |  |
| --- | --- | --- | --- | --- | --- | --- | --- | --- | --- |
| cluster | cluster | cluster | cluster | peak | peak | peak | peak | peak |  |
| p(FWE-corr) | p(FDR-corr) | equivk | p(unc) | p(FWE-corr) | p(FDR-corr) | T | equivZ | p(unc) | x,y,z {mm} |
| 0.969 | 0.777 | 54 | 0.027 | 0.22 | 0.274 | 5.11 | 4.68 | 0 | 22 -20 58 |
| 0.051 | 0.048 | 172 | 0 | 0.999 | 0.909 | 4.01 | 3.78 | 0 | 20 -4 52 |
|  |  |  |  | 1 | 0.909 | 3.59 | 3.42 | 0 | 12 -6 54 |
|  |  |  |  | 1 | 0.909 | 3.48 | 3.32 | 0 | 30 -2 58 |
| 0.723 | 0.581 | 78 | 0.01 | 1 | 0.909 | 3.8 | 3.6 | 0 | -20 0 46 |
|  |  |  |  | 1 | 0.909 | 3.68 | 3.5 | 0 | -30 0 50 |
|  |  |  |  | 1 | 0.971 | 3.21 | 3.09 | 0.001 | -26 -6 44 |

**Table S5.**
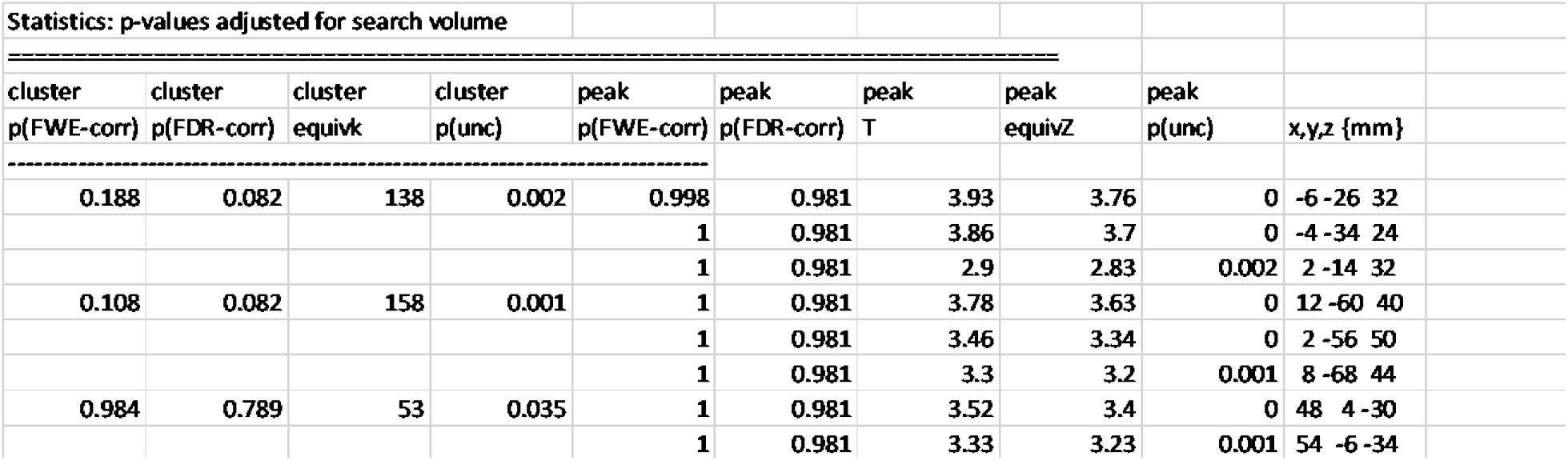
Whole-brain rAMYG seed-based connectivity with CAPS-Anxious Arousal symptom cluster severity. p < 0.005 (uncorrected height threshold), k = 50 arbitrary cluster threshold.

**Table S6.** Whole-brain rAMYG seed-based connectivity with CAPS-Startle Response symptom severity. p < 0.005 (uncorrected height threshold), k = 50 arbitrary cluster threshold.

| Statistics: p-values adjusted for search volume |  |  |  |  |  |  |  |  |  |
| --- | --- | --- | --- | --- | --- | --- | --- | --- | --- |
| cluster | cluster | cluster | cluster | peak | peak | peak | peak | peak |  |
| p(FWE-corr) | p(FDR-corr) | equivk | p(unc) | p(FWE-corr) | p(FDR-corr) | T | equivZ | p(unc) | x,y,z {mm} |
| 0.698 | 0.384 | 85 | 0.01 | 0.74 | 0.995 | 4.45 | 4.22 | 0 | -6 -38 46 |
| 0.022 | 0.021 | 218 | 0 | 0.976 | 0.995 | 4.11 | 3.93 | 0 | -126 |
|  |  |  |  | 1 | 0.995 | 3.1 | 3.01 | 0.001 | -28 -84 8 |
|  |  |  |  | 1 | 0.995 | 2.98 | 2.9 | 0.002 | -116 |
| 0.928 | 0.549 | 64 | 0.023 | 1 | 0.995 | 3.76 | 3.61 | 0 | -4 -34 24 |
| 0.291 | 0.166 | 122 | 0.003 | 1 | 0.995 | 3.7 | 3.56 | 0 | 6 -62 22 |
|  |  |  |  | 1 | 0.995 | 3.37 | 3.26 | 0.001 | 4 -62 34 |
|  |  |  |  | 1 | 0.995 | 3.13 | 3.04 | 0.001 | 6 -68 42 |
| 0.942 | 0.549 | 62 | 0.025 | 1 | 0.995 | 3.7 | 3.56 | 0 | 36 -84 2 |

